# A practical approach to curate clonal hematopoiesis of indeterminate potential in human genetic datasets

**DOI:** 10.1101/2022.10.21.22281368

**Authors:** Caitlyn Vlasschaert, Taralynn Mack, J. Brett Heimlich, Abhishek Niroula, Mesbah Uddin, Joshua Weinstock, Brian Sharber, Alexander J. Silver, Yaomin Xu, Michael Savona, Christopher Gibson, Matthew B. Lanktree, Michael J. Rauh, Benjamin L. Ebert, Pradeep Natarajan, Siddhartha Jaiswal, Alexander G. Bick

## Abstract

Clonal hematopoiesis of indeterminate potential (CHIP) is a common form of age-related somatic mosaicism that is associated with significant morbidity and mortality. CHIP mutations can be identified in peripheral blood samples sequenced using approaches that cover the whole genome, whole exome or targeted genetic regions; however, differentiating true CHIP mutations from sequencing artifacts and germline variants is a considerable bioinformatic challenge. We present a stepwise method that combines filtering based on sequencing metrics, variant annotation, and novel population-based associations to increase the accuracy of CHIP calls. We apply this approach to ascertain CHIP in ∼550,000 individuals in the UK Biobank complete whole exome cohort and the All of Us Research Program initial whole genome release cohort. CHIP ascertainment on this scale unmasks recurrent artifactual variants and highlights the importance of specialized filtering approaches for several genes including *TET2* and *ASXL1*. We show how small changes in filtering parameters can considerably increase CHIP misclassification and reduce the effect size of epidemiological associations. Our high-fidelity call set refines prior population-based associations of CHIP with incident outcomes. For example, the annualized incidence of myeloid malignancy in individuals with small CHIP clones is 0.03%/year, which increases to 0.5%/year amongst individuals with very large CHIP clones. We also find a significantly lower prevalence of CHIP in individuals of self-reported Latino or Hispanic ethnicity in All of Us, highlighting the importance of including diverse populations. The standardization of CHIP calling will increase the fidelity of CHIP epidemiological work and is required for clinical CHIP diagnostic assays.

## Introduction

Somatic mosaicism occurs across tissues as the human body ages.^1^ One of the best characterized examples of this is clonal hematopoiesis of indeterminate potential (CHIP), where a somatic mutation within a hematopoietic stem cell (HSC) leads to clonal production of myeloid lineage blood cells. The genes affected in CHIP are the same genes that drive myeloid neoplasms (e.g., *DNMT3A, TET2, ASXL1, JAK2, TP53*), though only 0.5-1% of CHIP cases progress to overt cancer per year.^2,3^ CHIP is associated with an estimated 40% increased risk of mortality^4^, largely due to disease beyond the hematopoietic system. An excess disease burden across multiple organ systems is seen among those with CHIP, including cardiovascular disease^5–8^, pulmonary disease and infections^9,10^, worsening kidney function^11,12^, osteoporosis^13^, and other inflammatory conditions.^14,15^ This systemic risk profile matches that which is observed in patients with chronic myeloid neoplasms.^16^ Dysregulated myeloid cell inflammatory activity appears to be a common mediator of CHIP-related organ damage across driver genes.^6,13,17–19^ CHIP driver mutations therefore appear to disrupt both HSC and daughter cell functions: they confer a competitive growth advantage to HSCs in order to achieve clonality^20^ and they disrupt the function of circulating myeloid cells to cause organ damage.

Somatic CHIP variants can be detected using genetic sequencing. A highly cost-effective and popular approach is to repurpose existing data from whole genome and whole exome sequencing conducted on peripheral blood cells of large cohorts for CHIP analyses. This method has been used to uncover the bulk of known CHIP disease associations in the population. In contrast, targeted sequencing approaches such as gene panels are frequently used in smaller cohort studies of CHIP due to their relative cost-effectiveness and greater sequencing depth, which increases accuracy.^21–26^ A CHIP mutation is currently defined as a driver variant identified in 2% or more sequencing reads (representing 4% of circulating diploid blood cells for heterozygous mutations)^3^, though targeted and error-corrected sequencing have revealed that clones below the 2% threshold are common.^27^ Conversely, genomes and exomes have limited sensitivity to detect small clones due to their modest sequencing depth.^28^ For this reason, estimates of CHIP prevalence and effect sizes for disease associations largely depend on the sequencing method used.^29^

Variant interpretation is another source of variability in CHIP ascertainment. Defining filtering parameters that permit accurate differentiation of somatic CHIP variants from sequencing artifacts, passenger mutations, and germline variants remains a challenge. Pre-specified mutation lists can help identify driver mutations.^4^ These lists identify missense variant sites as well as genes in which truncating or splicing variants may represent CHIP based on reported frequencies in myeloid cancers. However, additional filtering is usually required to remove false positives, such as sequencing artifacts masquerading as pathological variants. For example, a common *bona fide ASXL1* truncating variant is also a common sequencing artifact^30^ – seemingly since both DNA replication enzymes and DNA sequencing machinery have trouble handling this homopolymer section.^31^ Certain CHIP missense mutation hotspots have also been reported in germline DNA. This includes *DNMT3A* R882H, which causes a recognized heritable disorder Tatton-Brown-Rahman Syndrome^32^. Although error-corrected sequencing of blood or sequencing of a matched solid tissue sample can help differentiate CHIP from germline variation or sequencing artifact, these options are often not feasible for large cohorts. Conversely, large-scale human sequencing data and its linked demographic data can be leveraged to systematically identify erroneous CHIP calls and generate appropriate filtering criteria, such as a list of recurrent false positives.

In this work, we provide a generalizable framework for optimizing CHIP identification across any dataset. As an illustrative example, we apply this to the 454,787-person UK Biobank whole exome dataset and to the 98,560-person All of Us whole genome dataset and make these CHIP variant calls available for use by the global research community.

## Cohort descriptions

The UK Biobank (UKB) whole exome cohort is comprised of 454,787 individuals aged 40-70 at enrollment, when DNA was collected for sequencing.^33^ Participants were enrolled from 2008-2010 and were administered questionnaires, physical measurements, laboratory tests, and medical imaging at specified baseline and follow-up timepoints.^34^ As well, incident health outcomes since enrollment are tracked from hospitalization and general practice health records, as well as death and cancer registries. Whole exome sequencing (WES) was performed in two tranches: the first 50k using Illumina NovaSeq S2 flow cell and the second tranche of samples with the S4 flow cell with 75 base pair paired end reads and a modified version of the Integrated DNA Technologies xGen Exome Research Panel v.1.0, with ∼96% of targeted bases covered at a depth of 20x or greater and a median sequencing depth of ∼40x across sites^35^. The median age at enrollment for this cohort is 58 years old [interquartile range (IQR): 50-63]. CHIP has been ascertained by multiple groups in tranches of this data in the past using highly variable filtering criteria, resulting in differences in CHIP variant identification and prevalence estimates.^18,36,37^

The All of Us Research Program is an ongoing observational cohort study that is recruiting adult participants representative of the diversity of the US population.^38^ Linked health outcome data is pulled from participant survey questionnaires and linked electronic health record information. Over 536,000 participants of the targeted 1 million have been recruited to date and read-level whole genome sequencing (WGS) data for an initial tranche of 98,560 participants was released in June 2022. WGS was performed using Illumina PCR-free whole genome technology and sequenced on the NovaSeq platform with 150 base pair paired end reads to a median sequencing depth of 40x.^39^ Importantly, significant emphasis was placed in the development of the All of Us WGS to ensuring sequencing met clinical grade quality control specifications.^40^ Participants were enrolled in 2018-2021, and the median age at enrollment for this subset was 53 years old [IQR: 37-65]. CHIP has not previously been ascertained in this cohort.

### Putative somatic variant detection

The identification of somatic variants comprises two major steps: putative variant identification and variant filtering (Figure 1). In the first step, a somatic variant calling pipeline is used to scan aligned sequencing files for putative somatic variants. The most commonly used somatic variant calling pipeline to detect CHIP in the research setting is Mutect2, a package within the Genome Analysis ToolKit (GATK).^41^ Mutect2 uses local haplotype assembly and Bayesian modelling to detect single nucleotide alterations and small indels. Mutect2 can be used for whole genome, whole exome, or targeted sequencing data and is optimized for Illumina-based sequencing. For other sequencing platforms, a different variant caller may be necessary (e.g., TorrentVariantCaller for IonTorrent data^42^). Other commonly used somatic variant callers include Strelka^43^, VarDict^44^, VarScan^45^, and Shearwater.^46^

**Figure 1.**
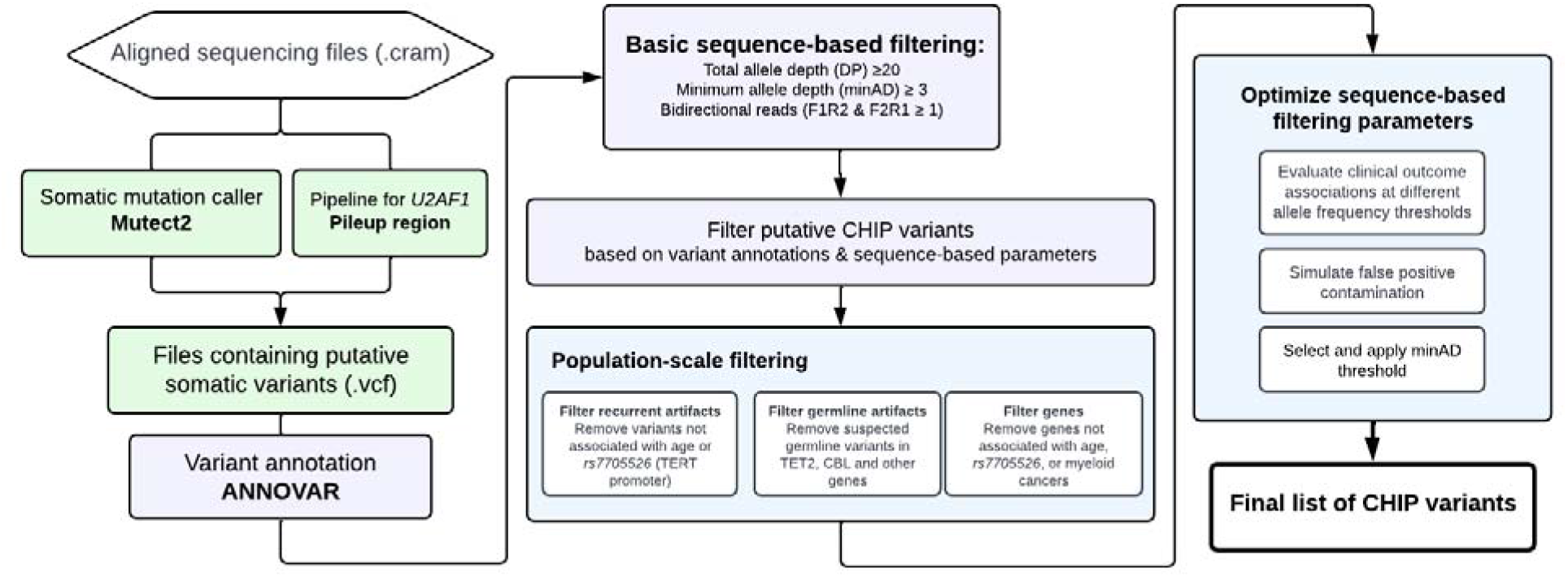
Schematic of CHIP variant ascertainment workflow. Putative somatic mutations are first identified using a somatic mutation caller and annotated for gene and protein level changes. Variants are then filtered based on an initial liberal set of parameters and filtered based on gene specific CHIP variant rules. In some genes all loss of function mutations are considered putative CHIP variants, whereas in other genes only specific missense mutations are included. Leveraging available large-scale sequencing data, we apply three filters to identify artifactual genes and variants. We then optimize the sequencing-based filtering parameters yielding a final CHIP mutation callset.

The foundational CHIP epidemiology papers describing CHIP identified a list of variants within 74 driver genes based established variant calling from the myeloid malignancy field.^47^ This list specifies candidate missense and indel variants for each gene, as well as a list of genes in which truncating and splice site variants might be considered CHIP (Supplemental Table 1). We limit our scan for putative CHIP variants to those contained in this list. We refer to variants in this list as the canonical CHIP driver variants, to differentiate them from more recently described gene variants observed to exhibit hematologic clonality but whose clinical consequences are less well defined.^22,48,49^ Of note, it is important to specify the transcript when scanning for variants as reference transcripts are periodically updated. For example, an *ETNK1* hotspot mutation that was located at N244 is now in position N155 (Supplementary Table 1).

We used Mutect2 to identify putative somatic variants in 73 of the 74 canonical CHIP driver genes in the UK Biobank and All of Us cohort aligned sequencing (CRAM) files. Altogether, 550,782 variants (average 1.21 variants per person) were output by Mutect2 for the UKB samples and 104,649 variants (average 1.06 variants per person) for All of Us samples. Variant calling pipelines such as Mutect2 cannot reliably identify variants in *U2AF1* in sequencing data that are mapped to the human GRCh38 (hg38) reference genome due to an erroneous duplication of the *U2AF1* locus on chromosome 21.^50^ A custom script was used to identify variants in *U2AF1*. The *pileup region* script counts mutated alleles present in reads that mapped to *U2AF1* in one of the two genomic loci, and variants corresponding to pre-specified hotspot locations are considered putative CHIP variants (Supplemental Table 1). For samples where reads were mapped to both loci, these were harmonized into one call and the allelic depth was taken as the average across both sites. This yielded 65,901 *U2AF1* variants in the UKB and 39,182 *U2AF1* variants in All of Us, for a total of 616,683 and 143,831 putative variants, respectively. ANNOVAR was then used to annotate all putative variants for downstream filtering.^51^

### Sequencing depth-based filtering

We first apply basic filtering to remove variants with low sequencing coverage: we removed variants with total read depth (DP) of less than 20, variants with minimum read depth for the alternate allele (minAD) of less than 3, and variants lacking variant support in both forward and reverse sequencing reads. We also removed variants below the 2% variant allele frequency (VAF) threshold conventionally used to define CHIP.^3^ This reduced the number of putative CHIP variants to 97,696 for UKB (0.21 variants per person) and 6,308 for All of Us (0.06 variants per person). It may be appropriate to relax or impose further stringency of these basic filtering criteria as we discuss later.

In large datasets, Mutect2 will output many multi-allelic variants (i.e., GT = 0/1/2 or 0/1/2/3). As these are difficult to interpret and might reflect artifactual variants, many groups opt to exclude these variants from further analysis. However, some *bona fide* biallelic variants may appear in the multiallelic variant list; for example, more than 300 *DNMT3A* P904L hotspot variants in UKB sequencing data were misclassified as multiallelic due to the artefactual imposition of a third allele with 0 reads at this site. Therefore, we recommend exercising caution and examining multi-allelic variants separately.

### Identification of false positives: sequencing artifacts

Among the list of variants that remain after basic sequence depth-based filtering, there are true CHIP variants, germline variants, non-pathogenic somatic (passenger) variants, and sequencing artifacts. We use a multi-step process to distinguish CHIP from these false positives, including filters that leverage the size of the UKB and All of Us datasets to identify recurrent false positives.

Variants that are present in ≥ 20 individuals in the UKB (0.04%) and in ≥ 15 individuals in All of Us (0.02%) were assessed for their potential to represent recurrent sequencing artifacts. For this, the association of each variant group with two established strong correlates of CHIP – age^2,4^ and a common genetic variant in the *TERT* promoter (*rs7705526*)^28,37,52^ – was tested. Variants that were not associated with either age or *rs7705526* at even a suggestive significance of p < 0.10 were removed from the dataset as they were suspected to represent sequencing artifacts. *DNMT3A* variants were exempt from this analysis as they are recognized to appear earlier in life and have decreased fitness with age^53^, and thus may not all be positively associated with age in this cohort of individuals aged 40-70.

This filtering strategy proved to be particularly useful for *ASXL1* variants, wherein 30 groups of truncating variants in exons 5 and 6 are reported in ≥ 20 people across both cohorts (Figure 2). Fifteen of the 30 variant groups examined were not associated with either age or *rs7705526*. We examined two putative variants that were present more than 2,500 times in the UKB dataset more carefully: *ASXL1* p.G646Wfs*12 and *ASXL1* p.G645Vfs*58. *ASXL1* p.G646Wfs*12 was initially thought to represent a mere sequencing artifact^54^, but has been more recently confirmed to be a *bona fide* mutation in some cases.^30,31^ Montes-Moreno *et al*. identified that sequencing methods that used PCR-only amplification introduced *in vitro* indels at the homopolymer locus encoding G645 and G646, whereas protocols using probe capture before PCR as well as Sanger sequencing did not.^31^ By comparing these sequencing methods, they identified that 9% of G646Wfs*12 variants in their dataset were true somatic variants, all of which had a VAF ≥ 10%, and that all G645Vfs*58 were artefactual. Given this, using a higher VAF threshold for *ASXL1* p.G646Wfs*12 and removing p.G645Vfs*58 variants has been suggested as a means to remove *in vitro* indel contaminants.^30,31^ We tested different VAF thresholds for both variants in UKB and similarly found that G646Wfs*12 variants with VAF ≥ 10% were strongly associated with age (Figure 2b), whereas no G645Vfs*58 variants VAF strata was associated with age (Figure 2c). Compared to rest of the UKB cohort, G646Wfs*12 variants with VAF ≥ 10% were associated with a 2.4-fold increased risk of death (HR 2.4, 1.9 to 3.0) and an 18-fold increased risk of incident myeloid cancer (HR 18.2, 95% CI 10.7 – 30.9) in Cox proportional hazards models adjusting for age, age^2^, sex, smoking status and 10 principal components of ancestry, further strengthening its credibility as a true CHIP variant. In All of Us, there were proportionally fewer artefactual G646Wfs*12 variants (lowest VAF 9.2%), and there were no G645Vfs*58 variants, likely due to a combination of the PCR-free sequencing library preparation methods and the longer sequencing read length. In total, there were 613 *ASXL1* p.G646Wfs*12 variants with VAF ≥ 10% in UKB and 119 in All of Us, making this among the top three most common CHIP variants in both cohorts alongside *DNMT3A* p.R882H and *DNMT3A* p.R882C (Supplemental Table 3). In many CHIP calling methods, frameshifts at homopolymer sites such as G646Wfs*12 are typically excluded.

**Figure 2.**
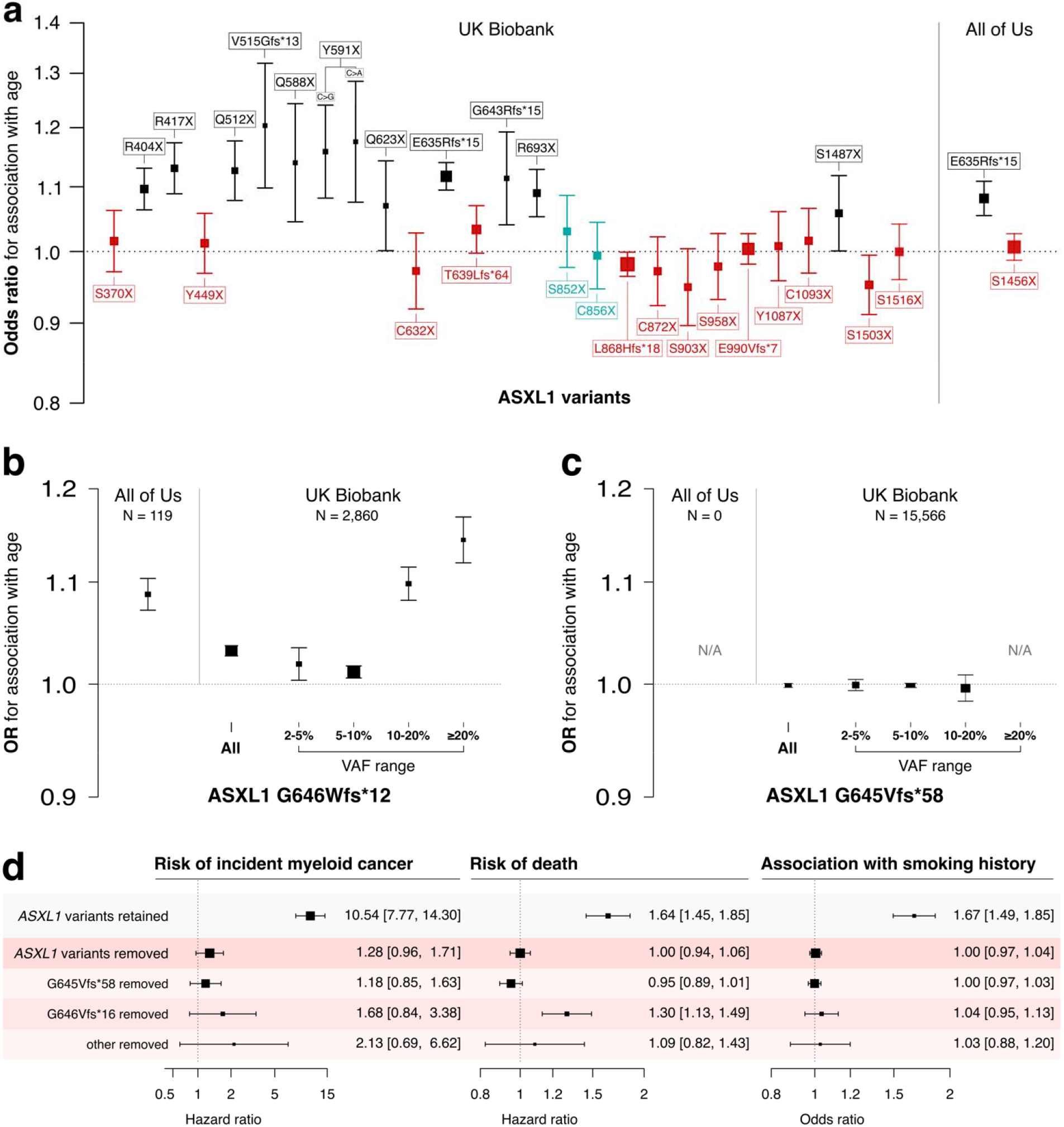
Verifying the association of common putative *ASXL1* variants with age can help distinguish true variants from recurrent artifacts. **a)** Association of all ASXL1 variants present ≥20 times in the UKB exome dataset and ≥15 times in the All of Us whole genome dataset. Variants not associated with age or a CHIP-associated *TERT* promoter variant (*rs7705526*) are colored in red. Variants associated with *rs7705526* only are coloured in blue. **b**,**c)** Association of *ASXL1* G646Wfs*12 and G645*58 with age across variant allele frequency (VAF) strata identifies specific large VAF subsets of G646Wfs*12 as somatic mutations while G645*58 appears to be an artifact of exome sequencing that is not present in All of Us. **d)** There is a significant association of *ASXL1* variants passing filtering with myeloid cancer, death and smoking, but minimal association with variants that were removed supporting that these removed variants are artifacts.

In total, we removed 146 variant groups in the UKB data that were not associated with either age or *rs7705526* (total: 37,953 variants) and 20 variant groups from the All of Us data (total: 681 variants), which are listed in Supplemental Table 2. We use this list to scan for potential artifacts from one dataset in the other dataset and removed 38 total variants from All of Us and 9 from the UKB dataset. These small numbers suggest that several artifacts are specific to the dataset being examined or to the sequencing methodology employed (Supplemental Table 2). All variants that were robustly associated with age and/or the *TERT* variant are indicated in Supplemental Table 3.

### Identification of false positives: germline variants

Variants disrupting the catalytic domains in TET2 and CBL are considered putative CHIP variants. In traditional CHIP calling methodologies, a binomial test is used to filter out possible germline variants in these domains; that is, a test to determine whether the measured read depth for the variant is statistically different from half of the sum of all sequencing reads at that site, as would be true for heterozygous germline variants. We conducted a binomial test across all *TET2* and *CBL* missense variants and flagged variants that failed the binomial test at p < 0.01. For many variant sites, there were multiple passing and non-passing variants, indicating that this site might exist as an acquired CHIP variant in some and as a germline variant in others. However, it is possible that some of the missense variants with a VAF near 50% represent large CHIP clones and not a germline variant. In an effort to recapture some of these large VAF clones in the UKB, we examined the association of all *TET2* missense variants with age and whether the addition of variants failing the binomial test improved or weakened the association with age (Supplemental Figure 1). We found that the addition of large VAF clones improved the association in 3 of the 9 examined sites – *TET2* H1904R, I1873T, and T1884A – suggesting that these are likely CHIP variants. These variants were exempt from the binomial test-based removal, including in All of Us.

We extended the binomial test examination to all other genes in both datasets in order to identify possible germline mutations therein. For variants present more than 3 times in either dataset, we examined the proportion of variants failing the binomial test. All variants in four variant groups – DNMT3A G298R, TP53 R110C, RUNX1 R223C and SUZ12 D725Vfs*18 (at the protein C-terminus) – failed the binomial test at p < 0.01; these 159 total variants were removed from the dataset. A list of variant groups where all variants failed the binomial test, i.e., recurrent germline variants, is in Supplemental Table 4.

### Sample quality verification

In addition to variant quality checks and rigorous filtering, it is important to verify sample quality. It is advisable to verify the number of variants per person and inspect variant lists for samples with an unusually high variant count, which could represent a poor-quality sample. For example, in All of Us, there were 10 individuals with 4 or more mutations. In 5 of these samples, all variants appeared to be of low quality and were likely artifactual, whereas the other 5 appeared to be credible. One example of each is presented in Supplemental Table 5.

### Revisiting the CHIP gene list

The originally defined CHIP gene list included 74 genes^5^; however, larger sample sizes enable us to prune this list. We found that putative variants in 16 of the 74 genes were not positively associated with either age nor observed in myeloid cancer cases in the either cohort, and not otherwise identified in published studies to drive myeloid CHIP (M-CHIP).^47^ These genes were: *SF3A1, GATA1, GATA3, PTEN, SF1, STAG1, IKZF2, IKZF3, PDSS2, LUC7L2, JAK1, JAK3, GNA13, KMT2A, KMT2D*, and *CSF1R* (Supplemental Table 6). Putative variants in these 16 genes were removed from the CHIP call set in both UKB and All of Us.

### Optimizing variant level allele depth thresholds

Altogether, 30,933 variants in the UKB exome cohort and 5719 variants in the All of Us first whole genome tranche passed the filtering steps. In the UKB, there were 7967 unique variants identified in this dataset, 180 of which were present ≥ 20 times. More than half of all variants were represented fewer than 20 times in the dataset (18,674 in total) and were mainly subject to the basic sequence-based filtering among the tests implemented above. In the initial filtering step, we set a relaxed minimum allele depth (minAD) threshold of 3 to increase sensitivity for CHIP variants. However, previous strategies to identify CHIP have used a minAD threshold as high as 6 in order to increase specificity. In order to identify minAD thresholds for UKB and All of Us, we first tested the associations between putative variants in minAD strata ranging from 3 to 6 with age and the *TERT* promoter variant (Figure 3). A minimum allele depth of 5 appeared optimal for UK Biobank exome samples compared to lower thresholds as the associations with age and the *TERT* promoter variant were maximized in this stratum. In order to estimate how much false positive misclassification might be present in the lower strata, we ran simulations where fractions of individuals with CHIP at minAD ≥ 5 were randomly replaced with CHIP-free individuals (Figure 3; see *Supplemental Methods* for details). The age- and *TERT* variant associations for minAD 3 stratum variants were smaller than the association seen when half of the minAD ≥ 5 group individuals were replaced at random, suggesting that approximately 50% of the variants in the minAD 3 stratum are expected to be false positives in the UKB (Figure 3a & b). This holds true for large variants within the minAD 3 stratum (VAF ≥10%), where the predicted contamination is approximately 40%. In All of Us, the predicted contamination was lower for minAD 3: it was estimated to be between 5-25% based on the age and rs7705526 association analyses (Figure 3c & d). In contrast to the UKB, each minAD stratum in All of Us appeared to capture distinct VAF ranges, and the association with age gradually increases across strata.

**Figure 3.**
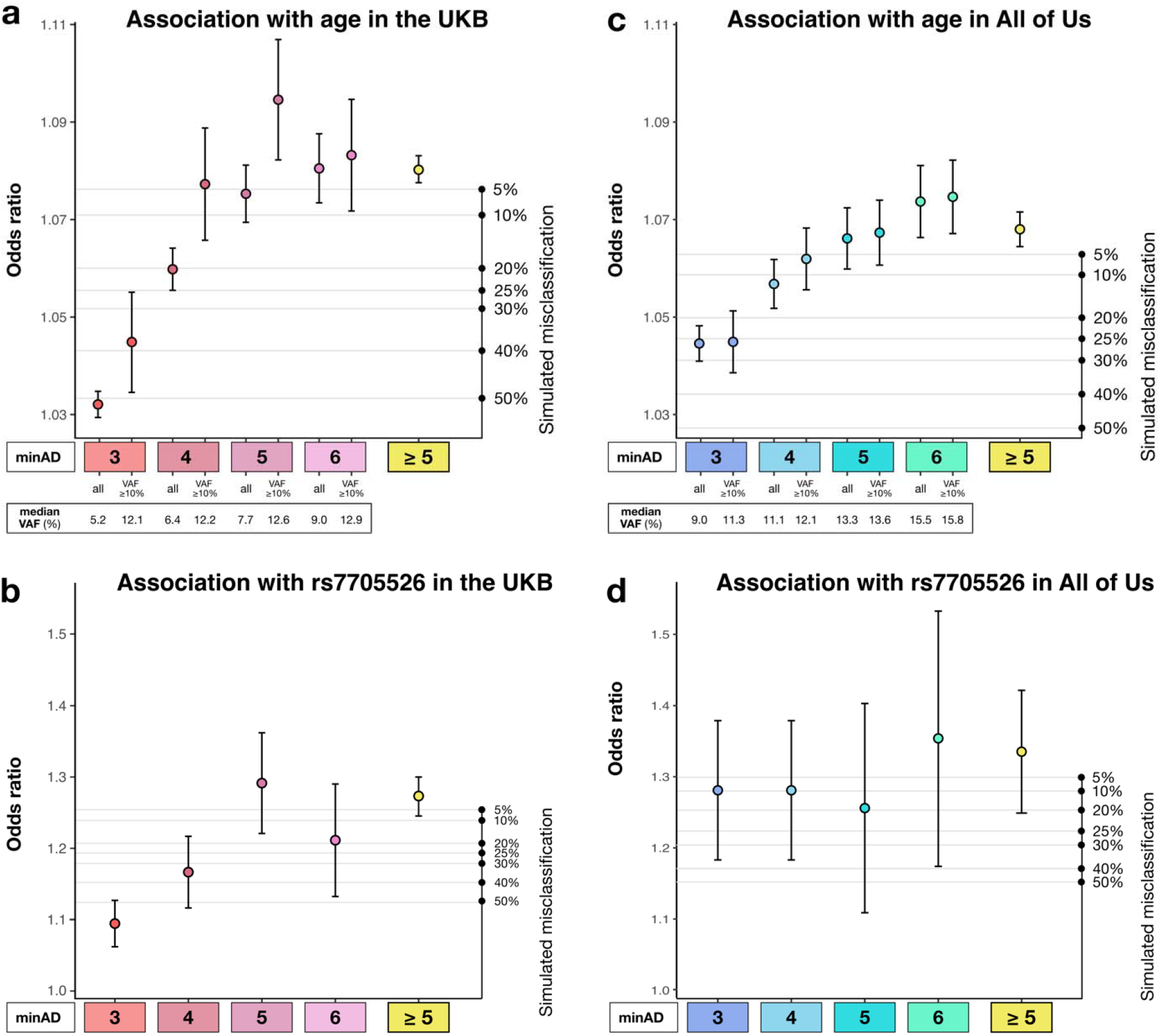
Association of CHIP variants defined by minimum allele depth (minAD) strata with age and *TERT* promoter variant rs7705526. Panels show the associations for strata of CHIP variants defined by minAD 3, 4, 5, and 6 with age (A,C) and the *rs7705526 TERT* promoter variant (B,D) in UK Biobank and All of Us Cohorts. The right axis plots the results of a simulation experiment wherein the specified proportion of samples in the minAD ≥ 5 CHIP dataset was randomly exchanged for individuals without CHIP in the dataset to estimate misclassification. Each simulation was run 20 times and the average result is shown.

Next, we tested how minAD thresholds of 3 and 5 estimated the CHIP-associated risk of death and incident myeloid cancer risk in the UKB. A minAD threshold of 3 underestimated the risk for both outcomes compared to previous population estimates^4^, including in subgroups comprised of CHIP hotspot mutations (present ≥ 20 times in the dataset) and non-hotspot mutations (present < 20 times; Figure 4). As lower minAD thresholds were predicted to contain significant contamination (Figure 3) and their inclusion did not improve incident outcome predictions (Supplemental Figure 2), only variants above a minAD threshold of 5 were included in our final set of UKB CHIP calls. In keeping with previous reports^2,4^, CHIP was associated with an 11-fold increased risk of incident myeloid cancer (HR 10.5, 95% CI: 9.1-12.1) and a 40% increased risk of death (HR 1.43, 95% CI: 1.36-1.50) in the UKB. Due to limited prospective data availability in All of Us (participants were enrolled in 2018-2021), incident event analyses were not performed in All of Us. Variants above a minAD threshold of 3 were included in the final set of All of Us CHIP calls.

**Figure 4.**
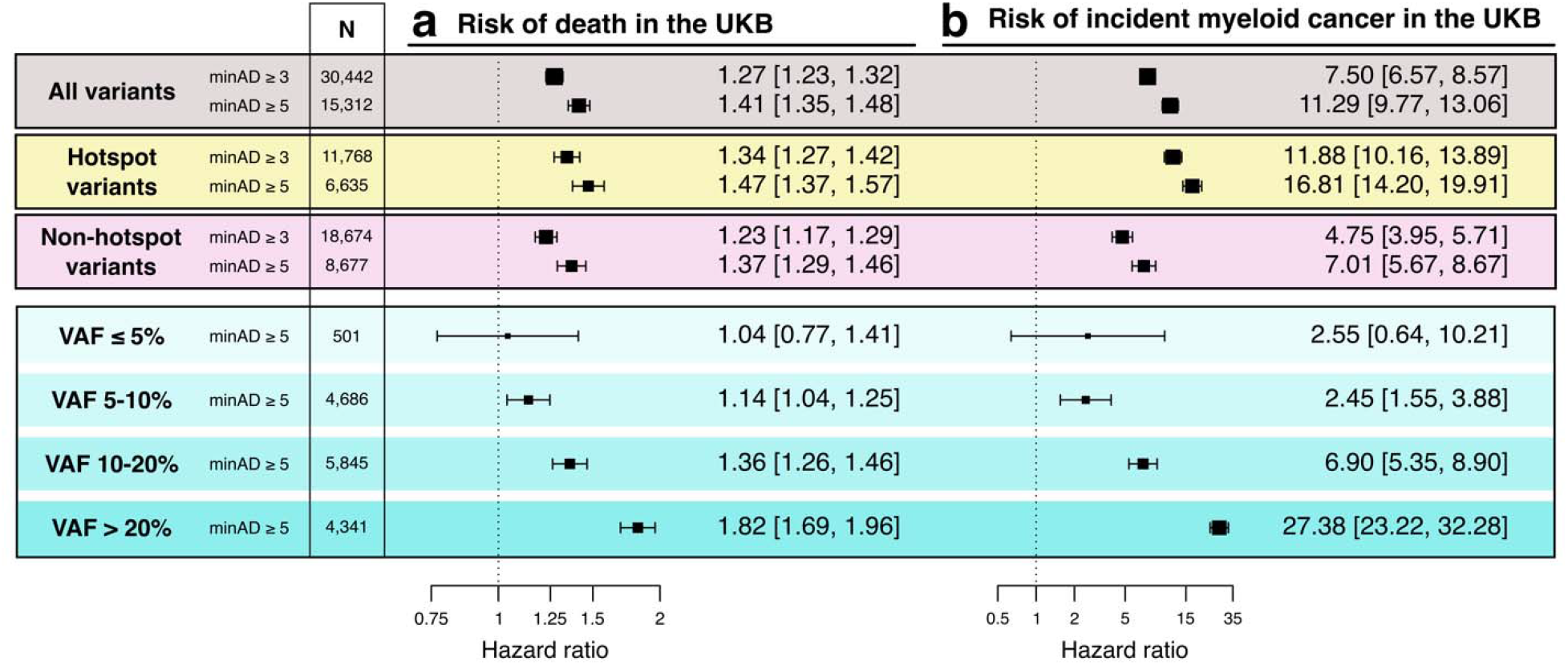
Associated risk of death and incident myeloid cancers with CHIP when defined using minimum sequencing allele depth (minAD) thresholds of 3 and 5 in the UK Biobank, assessed using Cox proportional hazards regressions adjusted for age, age^2^, sex, smoking history, and 10 principal components of genetic ancestry. The risk is greater for minAD ≥ 5, including when hotspot variants (defined as variants observed ≥ 20 times in the dataset) and non-hotspot variants (present < 20 times) are assessed separately. The risk increases proportional to the variant allele frequency (VAF) with nearly a ten-fold increase in risk between VAF <5% vs >20% strata.

### Profile of CHIP variants in the UKB and All of Us datasets

After exclusion of individuals with hematologic malignancies at baseline, there were 16,364 CHIP variants in 15,312 individuals in the UKB (3.4% prevalence) and 5195 CHIP variants among 4630 individuals in All of Us (4.8% prevalence). The distribution of genes affected are shown in Figure 5a and b. In both cohorts, the top three most common variants were DNMT3A R882H, DNMT3A R882C, and ASXL1 G646Wfs*12. CHIP was detected across all decades of age, and the prevalence rose sharply with age (Figure 5c). Other risk factors previously associated with CHIP prevalence that were replicated here included smoking history^36^ (OR 1.12, 95% 1.02-1.22) and self-reported Hispanic or Latino ethnicity^4,28^ (OR 0.82, 0.72-0.95; Figure 5d).

**Figure 5.**
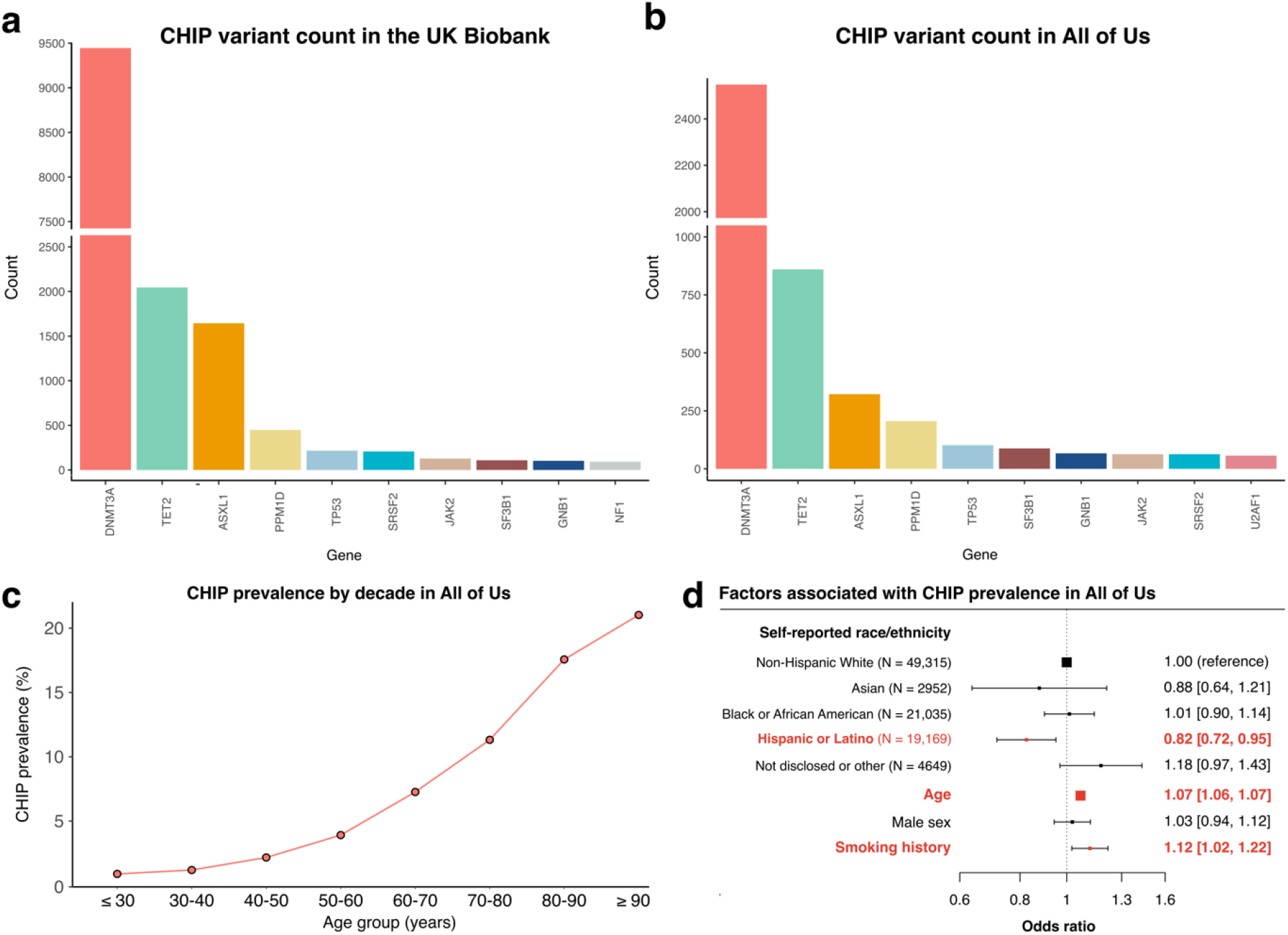
Profile of CHIP variants in the UK Biobank and All of Us. The distribution of genes affected by CHIP in (A) the UK Biobank and (B) All of Us are broadly similar. (C) CHIP prevalence increases with age in All of Us. (D) CHIP is associated with decreased prevalence in self-reported Hispanic or Latino ethnicity individuals and is positively associated with age and smoking history.

## Discussion

CHIP is an important contributor to human morbidity and mortality, though differentiating CHIP from sequencing artifacts and germline variants can be challenging in practice. We show how large datasets with paired genomic and demographic information can be leveraged to identify CHIP more accurately. This strategy builds upon methodologies used in the malignant hematology field to better understand blood cancer driver mutations. We provide a set of CHIP calls for the full UKB exome dataset and the All of Us interim 98k whole genome dataset, as well as a list of recurrent false positives encountered during variant interpretation. Our analysis was restricted to the originally described list of CHIP variants^4,5^, though could be adapted in the future to other genes have more recently been observed to exhibit clonality in blood cells.^47–49^

We highlight how the sequencing methodology can influence the types of artifacts detected. For example, methods that use PCR, such as exome capture-based sequencing, can introduce *in vitro* indel artifacts at the G645 and G646 homopolymer sites in *ASXL1*, whereas this was not seen with the PCR-free All of Us whole genome preparation method. In both the UKB and All of Us, *bona fide* G646Wfs*16 frameshifts were the most common *ASXL1* variant and were strongly associated with death and myeloid cancer risk, highlighting the importance of accurately identifying this variant. We also highlight how certain hotspot variants can be missing from datasets due to known issues with reference genome assemblies (i.e., *U2AF1* missing from both datasets due to hg38 error).

In CHIP variant interpretation, the minimum number of reads needed to support a somatic variant (minAD) is commonly set to 3 or greater. We show how small increases in this criterion can have large effects on risk estimates. Setting a higher minAD threshold may improve specificity in certain datasets, for example when sequencing involved PCR amplification steps. In contrast, a lower minAD threshold may be appropriate for datasets where fewer sequencing artifacts are expected, such as in All of Us whole genomes. In line with previous findings, we show that adverse outcomes of CHIP occur proportional to clone size. However, sequencing at 40x depth misses most small clones^28^, and deeper sequencing methods are needed to understand how small clones – including those below the 2% threshold – contribute to human disease on a population level.

## Data Availability

The UK Biobank calls and All of Us calls are in the process of being returned to the respective datasets and will be available to all registered researchers of the respective platforms upon publication.

## Data and code availability

UK Biobank calls and All of Us calls are in the process of being returned to the respective datasets and will be available to all registered researchers of the respective platforms upon publication. Scripts used to derive perform the various functions in this paper are available at: https://github.com/briansha/Cloud_Development/tree/master/DNANexus/Mutect2 (putative somatic variant identification), https://github.com/weinstockj/pileup_region (U2AF1 putative variant identification), https://github.com/briansha/Annovar_Whitelist_Filter_WDL (annotation and filtering pipeline).

## Acknowledgements and Funding

This work was supported by NIH Early Independence Award grant DP5 OD029586, a Burroughs Wellcome Fund Career Award for Medical Scientists, the E.P. Evans Foundation, RUNX1 Research Program, a Pew-Stewart Scholar for Cancer Research award, supported by the Pew Charitable Trusts and the Alexander and Margaret Stewart Trust and the Vanderbilt University Medical Center Brock Family Endowment and Young Ambassador Award to Dr. Bick.

C.V. receives financial support from the Canadian Institute of Health Research (CIHR) under the Canada Graduate Research Scholarship (RN410433–433120) and the Michael Smith Foreign Study Supplement (202106FSS-476208). A.J.S. received financial support from the US National Institutes of Health (NIH) under Ruth L. Kirschstein National Research Service Award F30DK127699 from the NIDDK. S.J. is supported by the Burroughs Wellcome Foundation Career Award for Medical Scientists, Foundation Leducq, Ludwig Center for Cancer Stem Cell Research, Leukemia and Lymphoma Society, the American Society of Hematology Scholar Award, and the NIH Director’s New Innovator Award (DP2-HL157540). M.S. receives funding from Leukemia and Lymphoma Society, the E.P. Evans Foundation, The Biff Ruttenberg Foundation, the Adventure Alle Fund, the Beverly and George Rawlings Directorship, and the NIH grants: 1R01CA262287, 1U01OH012271, P30 CA068485.

