## Supplemental Figures and Methods for "A practical approach to curate clonal hematopoiesis of indeterminate potential in human genetic datasets"

**Supplemental Material**

1. **Supplemental Figure 1** – Distribution of variant allele fractions (VAF) by age for *TET2* missense variant groups where only a portion of variants fail the binomial test.
2. **Supplemental Figure 2** – Association with death and incident myeloid cancers for CHIP defined by a minAD threshold of 3 compared to a minAD threshold of 5.
3. **Supplemental Methods**

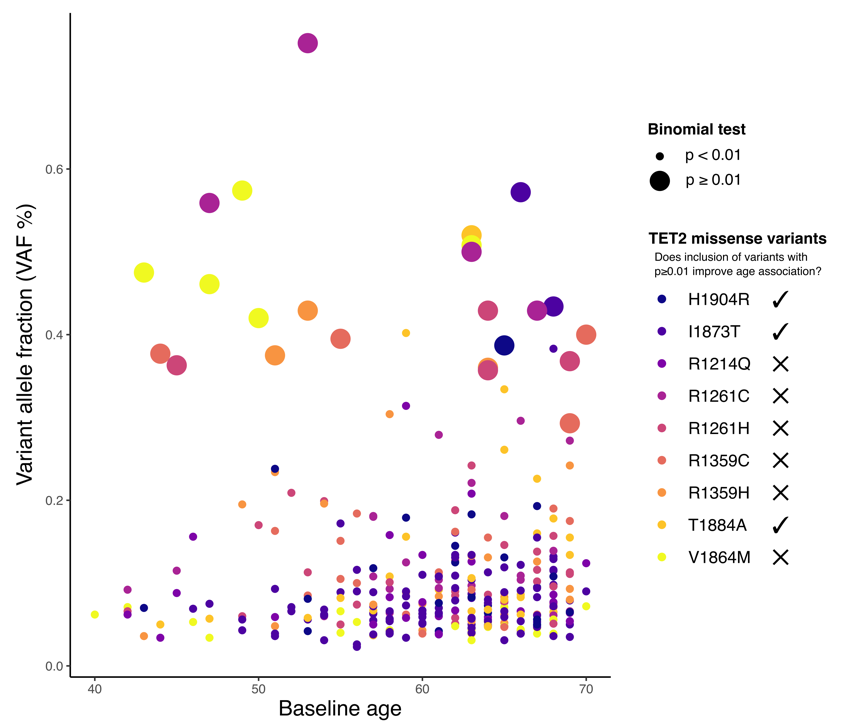

**Supplemental Figure 1.** Distribution of variant allele fractions (VAF) by age for *TET2* missense variant groups where only a portion of variants fail the binomial test. Cases where addition of variants failing the binomial test improves the association of VAF with age are indicated.

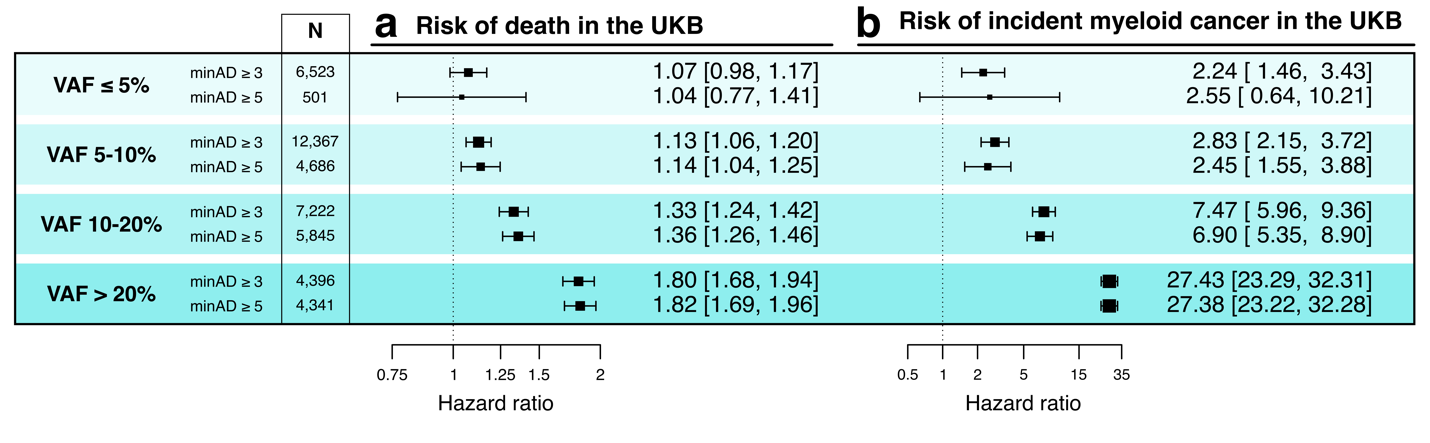

**Supplemental Figure 2.** Association with death and incident myeloid cancers for CHIP defined by a minAD threshold of 3 compared to a minAD threshold of 5.

**Supplemental Methods**

*Simulation testing*

We performed simulation testing in order to estimate the amount of variant misclassification present when using different minAD strata. We first identified that a minAD ≥ 5 had the greatest association with age and the *TERT* promoter variant in the UK Biobank. In our simulations, we replaced subsets of the CHIP call set defined by minAD ≥ 5 with randomly selected CHIP-free individuals from the cohort. For example, we replaced 5% of the dataset with CHIP-free individuals to estimate the effect that 5% of misclassification would have on the age- and *TERT* promoter variant associations. We carried out these simulations with 5%, 10%, 20%, 25%, 30%, 40% and 50% sample replacement, and each simulation was performed 20 times.
